# Psychological distress across adulthood: scale-equating in three British birth cohorts

**DOI:** 10.1101/2020.06.24.20138958

**Authors:** Hannah E. Jongsma, Vanessa G. Moulton, George B. Ploubidis, Emily Gilbert, Marcus Richards, Praveetha Patalay

**Author notes:** Hannah E. Jongsma is the corresponding author.

## Abstract

Valid and reliable life-course and cross-cohort comparisons of psychological distress are limited by differences in measures used. We aimed to examine adulthood distribution of symptoms and cross-cohort trends by scale-equating psychological distress measures administered in the 1946, 1958 and 1970 British birth cohorts. We used data from these three birth cohorts (N=32,242) and an independently recruited calibration sample (n=5,800) to inform the scale-equating. We used two approaches to scale-equating (equipercentile linking and multiple imputation) and two index-measures (General Health Questionnaire [GHQ]-12 and Malaise-9) to compare means, distributions and prevalence of distress across adulthood. While we consistently observed an inverse U-shape of distress across adulthood, we also observed measure and method differences in point estimates, particularly for cross-cohort comparisons. Sensitivity analysis suggested that multiple imputation yielded more accurate estimates than equipercentile linking. Though we observed an inverse-U shaped trajectory of psychological distress across adulthood, differences in point estimates between measures and methods did not allow for clear conclusions regarding between-cohort trends.

## Introduction

Despite the fact that common mental disorders are a leading cause of disease burden (Whiteford et al., 2013), with one in six adults in England meeting the threshold for a clinical diagnosis in 2014 (McManus et al., 2016), our ability to assess psychological distress reliably across time, person and place is limited. Limitations to comparability result from a lack of ‘gold standard’ measure and the plethora of instruments used, as well as from differences in mode of administration and response options.

### A life-course perspective

Psychological distress is a non-disorder based mental health outcome that includes symptoms of common mental health difficulties such as depression and anxiety. Life-course comparisons of psychological distress are crucial to understanding the distribution and determinants of as well as pathways to distress both within individuals and populations. In particular, the understanding of age-specific changes in the distribution of such distress, risk factors and aetiological mechanisms provides opportunities for targeted clinical and public health interventions at the appropriate timepoint in the life-course (Gondek et al., 2021).

Despite measurement limitations, there appears to be degree of consensus on the expected life-course distribution of psychological distress. In adulthood, the distribution of psychological distress is expected to follow an inverse-U shape with symptoms increasing from early adulthood to mid-life and then decreasing from mid-life to old age (Bell, 2014; Gondek et al., 2020; Spiers et al., 2012). Nonetheless, there is some evidence for other life-course distributions, including an increase in distress in the over-75s (Jokela et al., 2013). The limitations of relying on cross-sectional data across ages to determine the life-course shape of psychological distress are well recognised: mainly conflating of age and cohort effects (**Blanchflower & Oswald, 2000; Spiers et al., 2012; Thomson & Katikireddi, 2018**). One of the limitations to drawing stronger conclusions about the adulthood trajectory of symptoms in longitudinal data has been the use of different measures across time within the same population based cohort studies. For example, in the 1946 birth cohort the Present State Examination (PSE) is used at age 36, the Psychiatric Symptom Frequency (PSF) scale at age 43, and the General Health Questionnaire (GHQ -28) at ages 53, 63 and 69.

### Cross-cohort comparisons

Another area of great interest to population mental health researchers has been generational trends or cohort effects. Mapping such cohort effects aids prediction of population trends in healthcare usage as well as the social and wider burden of disease resulting from changes in population prevalence. If cohort differences are observed, it is important to understand the drivers of these to reduce negative or enhance positive impacts.

When trying to draw cross-cohort comparisons (do recent generations experience higher or lower levels of psychological distress than older generations?), similar problems around measurement arise - Comparisons are limited by different cohorts using disparate measures. On the rare occasion that identical measures have been used to compare cohorts and measurement equivalence has been formally established, there is some evidence that midlife psychological distress is higher in more recent mid-20^th^ century cohorts (Ploubidis et al., 2017). However, other studies based on creating pseudo-cohorts from repeated cross sectional data suggest different distributions with either more recent cohorts experiencing lower psychological distress (**Thomson & Katikireddi, 2018**) or U-shaped cohort effects with psychological distress highest in the oldest and most recently born cohorts in the United States (Keyes et al., 2014). To make reasonable comparisons across different life-stages where measures might not be consistent, understanding how scales compare and can be compared is an important prerequisite.

In addition to life-course and cross-cohort comparisons, there are several other uses when scales can be adequately equated. In mental health science, the use of disparate scales, even to measure the same construct, is widespread (Santor et al., 2006). For instance around 280 measures have been used over the last century to measure depression(Santor et al., 2006). Attempts to harmonise measures have taken several shapes (Farber et al., 2020), but all these approaches have their limitations, constraining the widespread use of common measures. Even if prospective harmonisation is successful, approaches to increase utility of historical data (retrospective harmonisation) are still warranted. Most recently there have been efforts from funders to mandate certain measures in all studies (Farber et al., 2020) to help create a ‘common language’ between these and any other measure used. The success of this approach to a great extent relies on scale equating being possible to a high enough quality. In this light, it becomes even more important to examine and test different approaches to scale equating and the conditions under which it might perform better or worse.

### Scale-equating approaches

The effects of variations in measurements on resulting estimates of the distribution and above-threshold prevalence of psychological distress as described above remain poorly understood and the empirical literature on scale-equating of mental health measures remains limited. Test- or scale-equating methods are more commonly used in the educational measurement literature to allow for various versions of a test to be used interchangeably or permit making valid comparisons between them (Chen et al., 2009; The Council of Chief State School Officers, 2018). Recently, various methods of scale-equating have been applied in the mental health field, in recognition of the great variety of instruments, response options and assessment modes available to capture common psychological symptoms. For example, applying Item Response Theory (IRT), studies have used General Partial Credit Modelling Fischer et al., 2011), as well as latent trait values and conversion tables (Fischer et al., 2012). Another study combined IRT-based approaches with equipercentile linking to cross-tabulate three legacy depression scales to a newly developed population-based depression metric (Choi et al., 2014). A further study applying an equipercentile linking approach used data from 14 clinical trials mapping the most commonly used observer-rated and the most commonly used self-reported measure of depression onto each other (Furukawa et al., 2019). To examine adolescent outcomes for children with mental health problems across three cohorts, Sellers and colleagues (2019) employed a multiple imputation approach using an externally recruited calibration sample, which involves imputing in the target scale for all participants using information from a sample that has both the target and original measure. A different approach that does not rely on external calibration samples compared subsets of harmonised items between scales, which limits analysis to the items or symptoms that are covered by the all the scales being compared e.g. an item on low mood (Gondek et al., 2020), Despite this recent interest in applying scale-equating methods to mental health measures, little is known about the effects of using these and other different approaches on resulting population-level estimates of distributions and above threshold prevalence of psychological distress, as no paper has previously compared different scale-equating methods.

### The present study

The availability of three successive national longitudinal cohort studies with mental health measures through adulthood (the 1946, 1958, 1970 birth cohorts) makes possible the investigation of both life-course progression in the same individuals longitudinally and cross-cohort comparisons if measures can be successfully equated or harmonised. The aims of the present study were two-fold: 1. to use two different scale-equating frameworks (equipercentile linking and multiple imputation) based on an external scale-equating (hereafter calibration) sample with two index measures (Malaise-9 and General Health Questionnaire [GHQ]-12) to equate all adult psychological distress measures in the three British birth cohorts; 2. to use the results of these different measures and methods to draw inferences on of the distribution and prevalence of psychological distress across adulthood and between cohorts.

## Methods

### Participants

We used data from four different datasets: a calibration sample, and the 1946, 1958 and 1970 British birth cohorts.

### Calibration sample

Our calibration sample (target *n*=5,000) was recruited independently from the birth cohorts between 7 June and 18 July 2019. We used quota-sampling to ensure at least 1,000 participants in each of five age groups (23-30, 31-40, 41-50, 51-60, 61-70) and representativeness of the general population in terms of sex, ethnicity, country of residence (England, Scotland, Wales, Northern Ireland) and highest level of education achieved. No further eligibility criteria were applied, and recruitment was halted when our target sample was reached for every age group. Participants also filled in the following questionnaires pertaining to wellbeing, which were not used in the present study but were part of a larger harmonisation effort (McElroy et al., 2020): the Center for Epidemiological Studies-Depression scale(Radloff, 2016), the Kessler Psychological Distress Scale(Kessler et al., 2003), the Mood and Feelings Questionnaire(Angold et al., 1995), the Short Form-36(Brazier et al., 1992), the Warwick-Edinburgh Mental Wellbeing Scale(Tennant et al., 2007) and the Office for National Statistics wellbeing questions(Office for National Statistics, 2018). Participants were compensated nominally for their time. Partially completed questionnaires were included.

### Cohorts

The MRC National Survey of Health and Development (1946 birth cohort) started as a maternity survey of 16,965 children born in one week in March in England, Scotland and Wales, then selected from this a social class-stratified sample of 5,362 individuals to follow over the life-course (Wadsworth et al., 2006). To date there have been 39 study sweeps (at birth, age 1, 2, 4, 6, 7, 8, 9, 10, 11, 13, 15, 16, 17, 18, 19, 20, 21, 24, 25, 26, 31, 36, 43, 47, 48, 50, 51, 52, 53, 54, 57, 59, 60-64, 68-70 years and a special Covid-19 sweep at age 74 (**MRC LHA & NSHD, 2022**)).

The National Child Development Study (1958 birth cohort) followed 17,415 children born in England, Scotland and Wales in a single week in March (**Power & Elliott, 2006**). There have been 13 sweeps of the full study population (at birth, age 7, 11, 16, 23, 33, 42, 44, 46, 50, 55 years and a special Covid-19 survey), as well as several sweeps in subsamples of the study (UCL Centre for Longitudinal Studies, 2022a).

The 1970 British Cohort Study started following 17,198 children born in England, Scotland and Wales in a single week in April (**Elliott & Shepherd, 2006**). To date there have been 15 study sweeps (at birth, age 22 months, 42 months, 5, 7, 10, 16, 21, 26, 30, 34, 38, 42 and 46 years), including some sweeps in subsamples of the study (UCL Centre for Longitudinal Studies, 2022b).

The analysis sample for the three birth cohorts was everyone for whom mental health data was available for at least one survey sweep in adulthood (18+ years).

### Main outcomes and measures

Our main outcome was psychological distress. In the calibration sample we collected data on all measures of psychological distress that were ever used in at least one of the three aforementioned birth cohorts (GHQ-12 and –28 (**Goldberg & Williams, 1988**) Malaise-9 (Rutter et al., 1970), Present State Examination [PSE] (Wing et al., 1967), Psychiatric Symptom Frequency Scale [PSF] (Lindelow et al., 1997); Supplemental Table 1) and administered them to the calibration sample. Following a counterbalanced design approach (Chen et al., 2009), order of questionnaire administration was randomised to prevent order effects.

We used the GHQ-12 and Malaise-9 as our index measures for both scale-equating methods. Our choice of index measures was based on their widespread use within the birth cohorts and beyond: the Malaise Inventory was widely used in UK population based cohorts and the GHQ-12 is used across a wider range of population-based and clinical studies. In addition the two measures have different response options (binary yes/no for Malaise and a 4-point Likert scale for the GHQ-12; hence resulting in differing ranges of possible scores and distributions). Using two index measures has the advantage of being able to assess robustness of the modelling procedure (each serve as the other’s sensitivity analysis). We collected and harmonised data on age in years, sex and highest level of education (none, GSCE or equivalent, A-levels or equivalent, degree or higher) to inform our multiple imputation models.

### Statistical analysis

We first composed descriptive statistics of all mental health measures collected in the calibration sample, and estimated correlations between these measures. In addition to multiple impuation, we used two approaches within an equipercentile linking framework: equipercentile linking and applying a calibrated cut-off. For our equipercentile linking approach to scale-equating, we cross-tabulated percentile rankings on the GHQ-12 or Malaise-9 with percentile rankings on remaining measures to determine equivalent scores (**Furukawa et al., 2019; Kolen & Brennan, 2014**). Within the equipercentile linking framework, we first identified a threshold score on the remaining measures most closely corresponding to that of the Malaise-9 (≥4) and the GHQ-12 (≥12). We applied this calibrated score back to the existing measures in the birth cohorts at each sweep to estimate the prevalence of mental distress. Throughout the paper, these results are described under the header ‘calibrated cut-off’. Secondly, using our equipercentile ranking, we converted scores on other measures in all sweeps of the birth cohorts to GHQ-12 or Malaise-9 and estimated means, variance and above-threshold prevalence of mental distress. These results are described as ‘equipercentile linking’. Details on how we meet the various assumptions associated with the equipercentile linking framework (**Kolen & Brennan, 2014**) are found in the Supplemental Methods.

Separately, we used a multiple imputation approach to scale-equating. Multiple imputation is a more robust extension of linear transformation-based scale-equating approaches (Lamprianou, 2007; Sellers et al., 2019), able to take account of stochastic error and uncertainty around single imputation transformed estimates. We coded data on covariates (age group, sex, level of education) and all psychological distress measures identically across all four datasets, with psychological distress measures coded at the scale-level per age-group. We appended the calibration sample to each of the cohort samples, separately for GHQ-12 and Malaise-9. Each dataset thus consisted of two samples, and (at least) two measures per age group. In at least one of these samples (the calibration sample) both measures were complete and data from this were used to impute values into the cohort sample (Supplemental Table 4). We used multiple imputation by fully-conditional specification using chained equations (**Little & Rubin, 2002**). We used the following generic algorithm: impute measure A based on measures B, C, sex, level of education, by age group. Analyses were conducted post-imputation, combining estimates across 50 imputed data sets using Rubin’s rule (White et al., 2011). Given the large percentage of missing data, this larger number was chosen to reduce variability from, and to improve robustness of, the imputation procedure. We estimated means, standard deviations and above-threshold prevalence of mental distress across adulthood in the cohorts.

### Sensitivity analysis

In both the 1958 (at age 42) and 1970 (at age 30) cohorts the GHQ-12 and Malaise-9 were jointly administered. We used this opportunity to assess comparability of prevalence yielding from the three methods described above to original estimates as an additional sensitivity analysis. For example, for the GHQ-12 calibration at age 30 in the 1970 cohort, we compared the prevalence yielding from the equipercentile linking, calibrated cut-off and multiple imputation approaches to the prevalence derived from the original GHQ-12 measure.

Throughout this paper we present results for the GHQ-12 in the main manuscript, with results for the Malaise-9 in the Supplemental Materials.

Our analysis plan was pre-registered on the Open Science Framework and can be accessed here: https://osf.io/7uc4j. We used Stata 16 for all our analyses (StataCorp, 2019).

### Ethics and consent

The authors assert that all procedures contributing to this work comply with the ethical standards of the relevant national and institutional committees on human experimentation and with the Helsinki Declaration of 1975, as revised in 2008. All procedures involving human subjects were approved by the UCL Institute of Education Research Ethics Committee (REC1210). Informed consent was obtained from all participants.

## Results

We recruited 5,800 participants into the calibration sample, distributed across five age groups (Supplemental Table 5). The analysis sample for the 1946 cohort consisted of 3,689 participants (68.7% of full cohort); for the 1958 cohort this was 14,814 (85.0% of full cohort); and for the 1970 cohort this was 13,739 (79.9% of full cohort). Missing data in the calibration sample was low (highest: 5.1% on GHQ-28, Supplemental Table 6). Means and standard deviations per questionnaire can be found in Supplemental Table 5. Correlations between measures varied between 0.68 (between Malaise-9 and GHQ-12) and 0.91 (between GHQ-12 and GHQ-28, Supplemental Table 7).

### Equipercentile linking

Scores calibrated to the GHQ-12 are detailed in Table 2, and calibrated means and standard deviations across the life-course in each of the three birth cohorts are detailed in Table 3 and Figure 1A. Psychological distress peaked in the 1946 birth cohort at age 43 (mean 8.54, standard deviation [SD]: 4.45) before declining to 5.20 (SD: 6.65) at age 69. In the 1958 birth cohort, psychological distress peaked at age 42 (mean: 7.26, SD: 6.23). In the 1970 birth cohort it peaked at age 26 (mean: 8.76, SD: 5.68). Across the life-course and across cohorts, distributions tended to be positively skewed (Supplemental Figure 1).

**Figure 1A:**
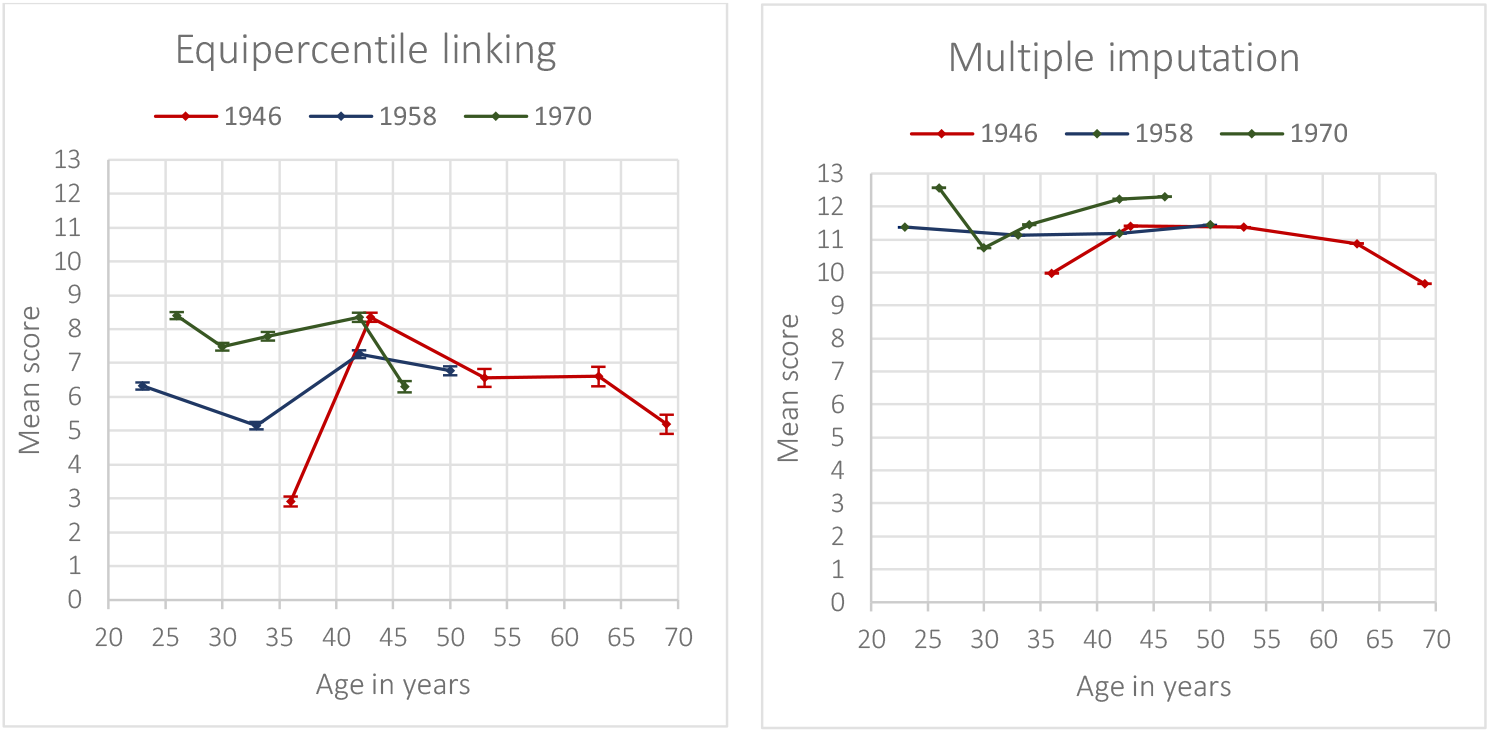
Mean scores and 95% confidence intervals derived for calibration against the GHQ-12.

**Figure 1B:**
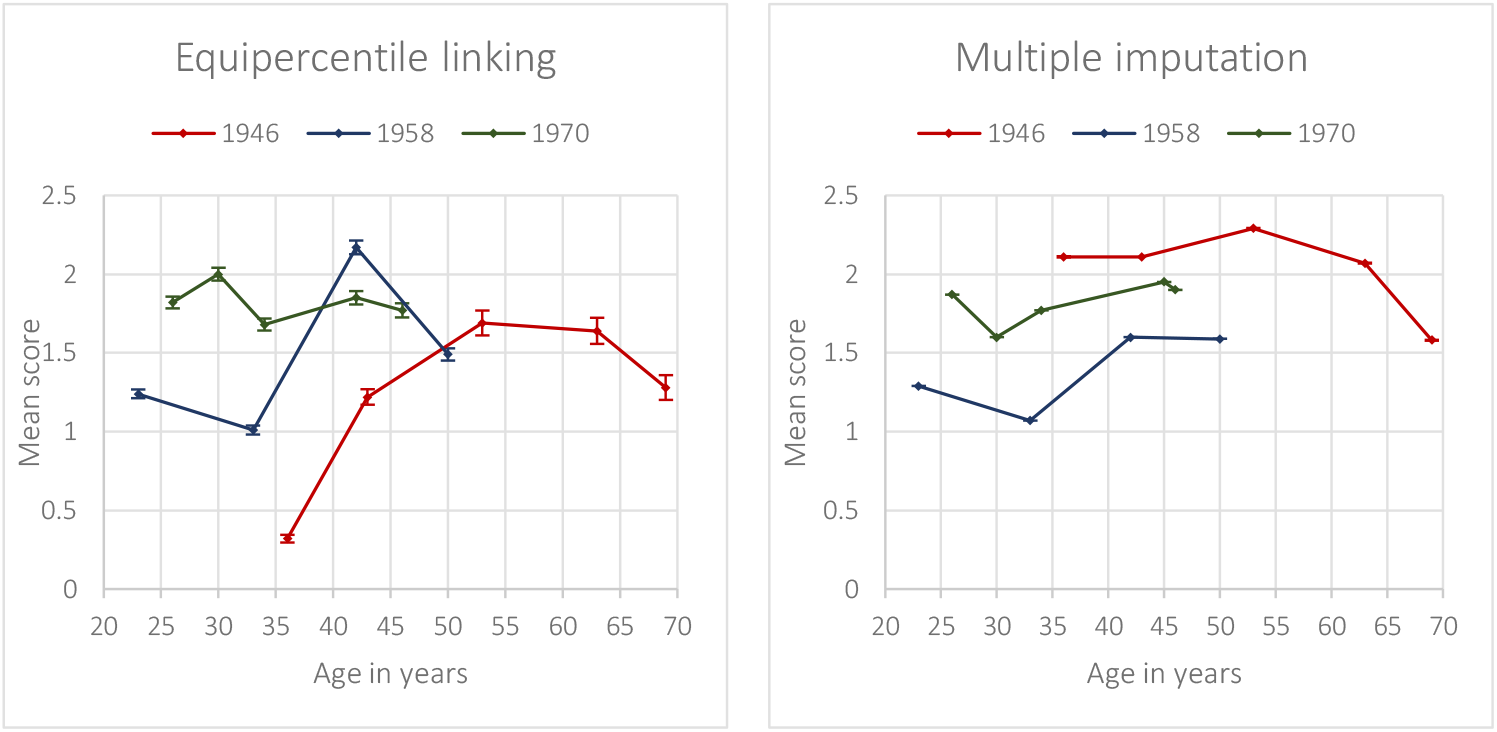
Mean scores and 95% confidence intervals derived for calibration against the Malaise-9.

### Calibrated cut-off

Corresponding calibrated threshold scores to the original threshold score of the GHQ-12 (12) were 2 for Malaise-9 and GHQ-28, 3 for Malaise-24 and PSE, and 11 for the PSF (Table 1). Distribution of above-threshold prevalence is detailed in Figure 2. For the calibrated cut-off scores, prevalence followed an inverse U-shape in the 1946 birth cohort, peaking at 35.9% at age 63, before declining to 27.3% at age 69. In the 1958 birth cohort, the shape was similar to the 1946 cohort and the peak was observed at age 42 (39.5%). Prevalence of psychological distress was relatively stable across the 1970 birth cohort, peaking at age 26 at 46.5%. Using the equipercentile linking method (where total scores were calibrated before the cut-off was applied), prevalence of psychological distress peaked in the 1946 birth cohort at age 63 at 35.9%, before declining to 27.3% at age 69. In the 1958 birth cohort, prevalence peaked at age 42 at 23.1%, and in the 1970 birth cohort the peak at age 42 was 28.7%.

**Table 1:** Calibrated scores and cut-offs against the GHQ-12.

| Calibrated equivalent scores per questionnaire |  |  |  |  |  |
| --- | --- | --- | --- | --- | --- |
| GHQ-12 score | Malaise-9 | Malaise-24 | GHQ-28 | PSE | PSF |
| 0-3 | 0 | 0 | 0 | 0 | 0 |
| 4 |  |  |  |  |  |
| 5 |  |  |  |  |  |
| 6 |  |  |  |  | 1,2 |
| 7 |  | 1 |  | 1 | 3-6 |
| 8 |  | 2 |  | 2 | 7-9 |
| 9 | 1 | 3 |  | 3 | 10,11,12 |
| 10 |  | 4 |  | 4 | 13-16 |
| 11 | **2** | 5 | 1 | 5 | 17,18 |
| **12** |  | **6** | **2,3** | **6** | 19-24* |
| 13 | 3 | 7,8 | 4,5 | 7 | 25-30 |
| 14 | 4 | 9 | 6 | 8 | 31-34 |
| 15 |  | 10 | 7,8 | 9 | 35-38 |
| 16 | 5 | 11 | 9,10 |  | 39-42 |
| 17 |  | 12 | 11 | 10 | 43,44 |
| 18 | 6 | 13 | 12,13 |  | 45-47 |
| 19 |  |  | 14 | 11 | 48-50 |
| 20 | 7 | 14 | 15 |  | 51 |
| 21 |  | 15 | 16,17 | 12 | 52-54 |
| 22 |  | 16 | 18 | 13 | 55-58 |
| 23 | 8 | 17 | 19 |  | 59-61 |
| 24 |  | 18 | 20,21 | 14 | 62-64 |
| 25 |  |  |  |  | 65,66 |
| 26 |  | 19 | 22,23 | 15 | 67-70 |
| 27 |  | 20 |  |  | 71,72 |
| 28 | 9 |  | 24 | 16 | 73-75 |
| 29 |  | 21 | 25 |  | 76,77 |
| 30 |  | 22,23 | 26 |  | 78-80 |
| 31,32 |  | 24 | 27 |  | 81-85 |
| 33-35 |  |  | 28 |  | 86-90 |

Equivalent cut-off scores are in **bold**
\* Equivalent cut-off score for PSF is 20

**Figure 2:**
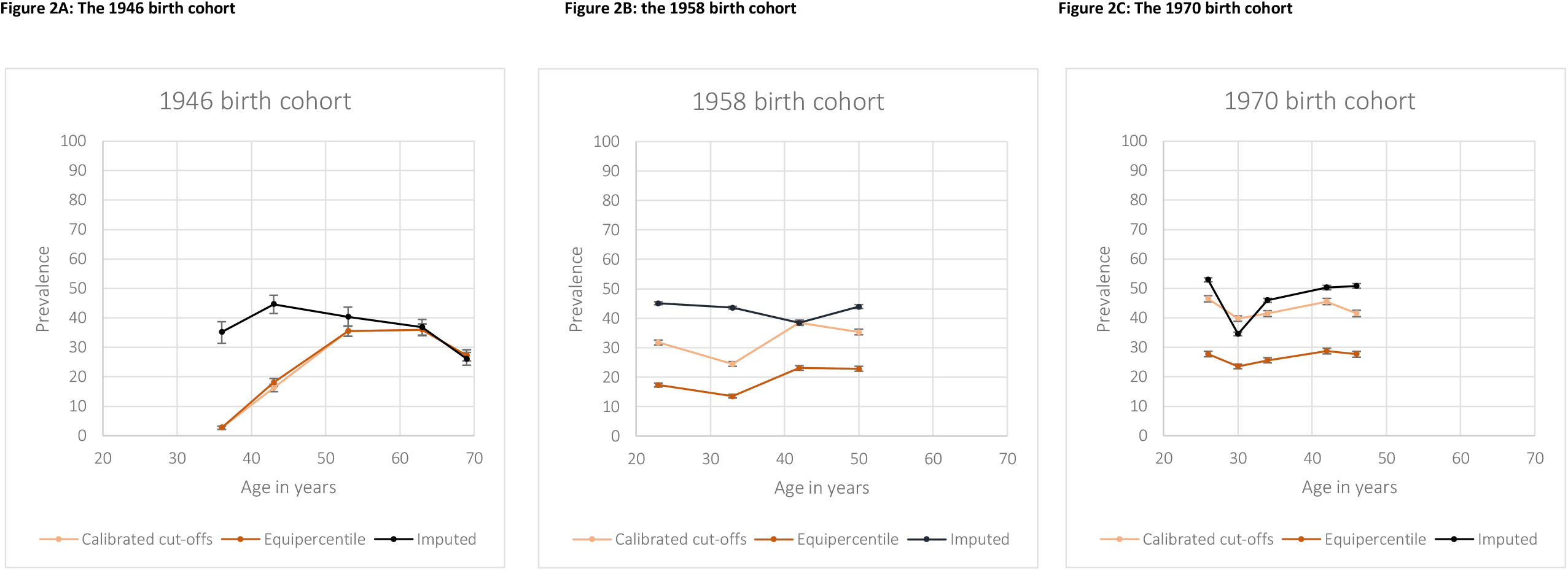
Prevalence of psychological distress and 95% confidence intervals across adulthood, calibrated against the GHQ-12.

### Multiple Imputation

Means and standard deviations of psychological distress across the life-course based on multiple imputation are detailed in Table 2 and Figure 1A. In the 1946 cohort, mean scores peaked at age 43 (mean 11.41, SD: 0.35) before declining to 9.66 (SD: 0.19) at age 69. In the 1958 and 1970 birth cohorts, means were approximately similar across the life-course, varying from 11.13 (SD: 0.35) at age 33 to 11.44 (SD: 0.20) at age 50 in the 1958 birth cohort and from 10.74 (SD: 0.04) at age 30 to 12.57 (SD: 0.42) at age 26 in the 1970 birth cohort.

**Table 2:** Means and standard deviations of measures calibrated against the GHQ-12 across adulthood.

| Cohort | Age | Multiple Imputation | Equipercntile linking | Original measure<br>(where available) |
| --- | --- | --- | --- | --- |
|  |  | Mean GHQ-12 (SD) | Mean GHQ-12 (SD) | Mean GHQ-12 (SD) |
| 1946 | 36 | 9.98 (0.38) | 2.91 (4.25) |  |
|  | 43 | 11.41 (0.35) | 5.35 (3.89) |  |
|  | 53 | 11.37 (0.22) | 6.56 (7.28) |  |
|  | 63 | 10.88 (0.19) | 6.60 (6.87) |  |
|  | 69 | 9.66 (0.19) | 5.19 (6.66) |  |
| 1958 | 23 | 11.37 (0.35) | 6.32 (5.97) |  |
|  | 33 | 11.13 (0.35) | 5.15 (5.97) |  |
|  | 42 | 11.19 (0.05) | 7.26 (6.23) | 11.06 (4.79) |
|  | 50 | 11.44 (0.20) | 6.77 (6.59) |  |
| 1970 | 26 | 12.57 (0.42) | 8.40 (4.93) |  |
|  | 30 | 10.74 (0.04) | 7.48 (6.11) | 10.67(4.52) |
|  | 34 | 11.45 (0.61) | 7.79 (6.33) |  |
|  | 42 | 12.23 (0.42) | 8.35 (6.37) |  |
|  | 46 | 12.30 (0.36) | 7.63 (6.86) |  |

Corresponding above-threshold prevalence estimates of psychological distress are detailed in Figure 2. The peak in the 1946 birth cohort occurred at age 43 at 44.6%, before declining to 36.8% at age 69. In the 1958 birth cohort, prevalence declined from a peak at age 23 (45.1%) until age 42 (38.5%) before increasing again at age 50 (44.0%). In the 1970 birth cohort, prevalence was highest at age 26 (52.9%) and lowest at age 30 (34.5%).

### Life-course mental health and cross-cohort comparisons

Although broad patterns in distribution and above threshold prevalence over adulthood appear similar across both index measures and the two scale-equating methods used in this paper, point estimates differ substantially. For instance, when examining the mean scores and standard deviations at age 36 in the 1946 birth cohort, a three-fold variance is observed for the GHQ calibration (2.91 [SD: 4.25] for the equipercentile linking method, and 9.98 [SD: 0.38] for the imputation approach). Prevalence scores for this sweep varied from 2.7% in the equipercentile linking and calibrated threshold approaches using the GHQ-12 to 35.2% using a multiple imputation approach.

Regarding cross-cohort comparisons of the prevalence of psychological distress calibrated against the GHQ-12, all approaches suggest that prevalence is highest in the 1970 birth cohort, though there is considerable variation in point estimates. Calibration against the Malaise-9 using a multiple imputation approach suggests that prevalence in the 1946 birth cohort is higher than in the other two cohorts, whereas using both a calibrated cut-off and equipercentile linking approach suggest that prevalence is comparable across the three cohorts.

### Comparison of the two index measures

Full results for the various methods of calibration against the Malaise-9 can be found in the Supplemental Results. Figure 1 details the mean scores across the adulthood sweeps in all three cohorts for both the equipercentile linking and multiple imputation methods, for both GHQ-12 (Figure 1A) and Malaise-9 (Figure 1B). There appear to be larger differences in means between both methods for the GHQ-12 compared with the Malaise-9, though this was not formally tested. Calibration against the GHQ-12 yielded higher prevalences than calibration against the Malaise-9 (Figure 2, Supplemental Figure 6).

Though broad patterns of psychological distress across the life-course were similar for GHQ-12 and Malaise-9 with prevalence highest in mid-life across the three cohorts, the curve was much flatter for the Malaise-9 across all methods (Supplemental Figure 6). Whereas using the GHQ-12 as an index measures seems to suggest slightly higher psychological distress in the younger cohort, using Malaise-9 suggests that psychological distress is lowest in the 1958 cohort.

### Sensitivity analysis

Additional sensitivity analyses resulting from comparing both calibration methods to original prevalence estimates at age 42 in the 1958 cohort and age 30 in the 1970 cohort suggested more accurate estimates using the multiple imputation method with both measures (Supplemental Table 10). When calibrating against the GHQ-12, the equipercentile linking method underestimated mean scores and prevalence in both cohorts. The multiple imputation method was close to original estimates in both cohorts (1958 original mean: 11.06 [SD: 4.79], multiple imputation 11.19 [SD:0.05] and 1970 original mean: 10.67 [SD:4.52], multiple imputation: 10.74 [SD:0.04]). The calibrated cut-off methods yielded relatively similar prevalence to the original estimate in the 1958 cohort (original prevalence: 36.1%, calibrated cut-off: 38.5%), but overestimated prevalence in the 1970 cohort (original prevalence: 32.9%, calibrated cut-off: 46.5%).

## Discussion

While broad patterns of psychological distress were similar across adulthood, the equipercentile linking scale-equating framework (both equipercentile linking and using a calibrated cut-off) yielded lower means and standard deviations across the life-course compared with the multiple imputation approach. While this held true across both index measures, differences appeared to be larger for the GHQ-12 than the Malaise-9. Cross-cohort comparisons were more susceptible to methodological effects. In general, using the GHQ-12 as an index measure yielded higher prevalence estimates than the Malaise-9. Sensitivity analyses using study sweeps with both index measures suggested that a multiple imputation approach leads to more accurate mean and prevalence estimates, whereas the equipercentile approaches yielded under- or over-estimates.

### Comparison with existing literature

The previously reported (Bell, 2014; Gondek et al., 2020; Spiers et al., 2012) inverse U-shape, across adulthood was observed in the 1946 birth cohort for calibration against the GHQ-12 and Malaise-9 for the calibrated-cut-off and equipercentile linking methods, though for both index measures the multiple imputation method showed a gradual decline in prevalence of psychological distress across the life-course. This pattern was less clearly observed in the 1958 and 1970 cohorts across both index measures and calibration methods used, likely due to the fact that these cohorts are still in middle age and prevalence of psychological distress has only started to decline marginally.

As per previous research comparing age 42 years sweep across the 1958 and 1970 cohorts (Gondek et al., 2020; Ploubidis et al., 2017), we find that, based on most methods and measures, the 1970 cohort has a higher prevalence of psychological distress at most ages compared to the 1958. However, we can draw no clear conclusions about any trends when also including the 1946 cohort. In the current study, we see lower prevalence in the 1946 cohort when using the GHQ-12 as index measure, but higher prevalence of psychological distress in this cohort when using the Malaise-9 as index measure. Previous studies in North America have found that some older and more recent cohorts have higher distress, demonstrating a U-shape in between-cohort effects (Keyes et al., 2014); and other UK-based studies have observed lower prevalence in more recent cohorts (**Thomson & Katikireddi, 2018**). It is important to note that the birth cohorts included in the present study were born across just 24 years in mid-20^th^ century Britain, and hence we cannot extrapolate findings to recent cohorts where higher distress is increasingly reported. Our sensitivity analysis using cohort sweeps where both GHQ-12 and Malaise-9 were administered could not provide insight into why clear conclusions about cross-cohort trends could not be drawn. Though it seems to suggest that multiple imputation might be more reliable than equipercentile linking. However, even just focussing on the multiple imputation findings we see prevalence of psychological distress increasing in more recent cohorts using the GHQ-12, yet lowest distress in the 1958 cohort using the Malaise-9.

### Strengths and limitations

These results should be interpreted in the light of the strengths and limitations inherent to this study. We used three nationally representative birth cohorts, and our calibration sample had sufficient coverage across the whole distribution of mental health and was broadly representative of the current general population of the United Kingdom in terms of age, sex, level of education and country of residence. Our study was methodologically robust and was designed to allow for assessing reliability across methods: we used two different index measures and scale-equating methods, enabling us to describe differences on the basis of these. This is in contrast to previous literature applying these methods which typically only use one method and measure (Choi et al., 2014; Fischer et al., 2011; Fischer et al., 2012; Sellers et al., 2019) resulting in limited evaluation of the reliability of any findings based on these approaches. Using an Item Response Theory (IRT) calibration model as a further methodology was unsuitable, as its key assumptions could not be met (Hambleton et al., 1991): The two questionnaires used as index measures do not have a common set of anchor items, nor do the remaining items have identical response options.

However, there are also some limitations inherent to our study design. As we only used data from the United Kingdom, we are uncertain about the generalisability of our results to an international context. Mode of questionnaire administration differed between our calibration sample (all self-reported online) and the cohort samples (either self-report via a paper questionnaire or interviewer-administered), and this might have led to higher reporting of mental health symptoms in the calibration sample (Epstein et al., 2001). Finally, we are utilising responses today to equate responses given at a previous point in time, as far back as the early 1980s. However, there appears to be no evidence that within-individuals and cross-cohort interpretation of the Malaise-9 changes over time (Ploubidis et al., 2019);. Also, measurement invariance analyses (Supplemental Table 3) indicate that younger and older respondents today answer the measures used in this study similarly, increasing confidence in the longitudinal comparisons made.

### Interpretation

While life-course patterns of psychological distress were similar across both index measures and scale-equating methods, point estimates were not. When comparing methodologies, the imputation method yielded higher means and standard deviations than the equipercentile linking method, and sensitivity analyses indicate the former might be the less biased approach in this scenario. While means are not directly comparable across index measures due to different scale ranges, prevalence estimates were higher using the GHQ-12.

These differences have little bearing on the longitudinal symptom profile (we confirmed an inverted U-shape over the life-course). There are two hypothetical explanations for this inverted U-shape: it might be artefactual because the instruments we use are poor at capturing important aspects of mental health in later life; or it might be a reflection of genuine better mental health in later life (either through a reduced perception of pressure through socioemotional selectivity or through eudaemonic processes) after a period of greater stress in mid-life (potentially reflecting the multiple stressors faced by many of childcare, career pressures and caring for elderly parents (Willis et al., 2010)). However, these differences do have substantial implications for cross-cohort comparisons. For instance, when examining means derived through the equipercentile method calibrated against the Malaise-9 (Figure 1), means are higher in the 1958 and 1970 cohort, and any mid-life peak is earlier in these cohorts. However, when using the same index measure and applying our multiple imputation-based approach, the mean is highest in the 1946 birth cohort, and there is no discernible mid-life peak in the other two cohorts. This method- and measure-dependency leaves us unable to make strong conclusions about whether more recent generations experience poorer mental health.

It is important to note that the Malaise-9 has less variance given the range from 0-9, compared to the GHQ-12 (range 0-36) and most of the other measures that we calibrated. We speculate that some of the discrepancies in the results we observe between these different measures might be due to differences in their scales (for instance a score of between 8 and 10 on the GHQ-12 gives a score of 1 on the Malaise-9, Supplemental Table 7). If this is indeed an important part of the consideration, then scale-equating between measures with similar ranges and variance is more likely to yield reliable estimates than measures with vastly different variances. This might also explain why the multiple imputation approach appears to be more reliable than the other two approaches, as it does not superimpose substantially larger or smaller variance. An important implication of our findings is the need for formal statistical simulation studies to investigate the conditions where different calibration and or harmonisation methods return unbiased results, especially if, as we have shown, bias occurs when measures with large discrepancy with respect to their range and standard deviation are calibrated.

This methodological and measure-based heterogeneity in point estimates across the life-course and between cohorts calls into question the robustness of papers using one index measure only to estimate mental health outcomes, and we would recommend where possible for studies using scale-equating approaches to use two index measures with similar range. On the basis of our analyses, it would seem that multiple imputation-based scale-equating methods are better suited to scenarios such as this, where an external calibration sample is used.

The potential inadequacy of these scale-equating approaches in translating findings from one measure to another has considerable implications with regards to recent announcements to mandate the use of certain measures in all research pertaining to certain common mental health symptoms (Farber et al., 2020). The arguments in support for this include being able to use these measures to act as a common language, via scale-equating approaches, so as to be able to draw comparable comparisons and conclusions across a range of studies in various settings including population studies, trials and routine clinical monitoring. The findings here highlight that these approaches are problematic even for robust population-level inferences; and individual-level inferences (where a score of X on this measure means a likely score of Y on the other measure for each individual) are likely to be even more error-prone (Piantadosi et al., 1988).

### Conclusion

We used three different scale-equating methods in two distinct frameworks and two different index measures to calibrate psychological distress measures used in three British birth cohorts against an independently recruited calibration sample. Our subsequently derived mean scores and above cut-off prevalences showed heterogeneity across both measure and method. While this had some implications for estimations of psychological distress across the life-course, we mainly observed an inverse-U shaped trajectory across adulthood. However, the method and measure had the most severe consequences for cross-cohort comparisons, where although there are indications that distress is higher in the 1970 compared to 1958 birth cohort, consistent trends across all three cohorts were not observed. We therefore should be cautious in interpreting calibration studies that have relied on one method and one index measure only. We recommend that future studies using scale-equating approaches to compare mental health across time-points or datasets use more than one measure to increase reliability of findings. We also highlight the need for formal statistical simulation studies to investigate the conditions where different calibration and or harmonisation methods return unbiased results.

## Data Availability

Data from the 1946 birth cohort is available to bona fide researchers upon request to the NSHD Data Sharing Committee via a standard application procedure. Further details can be found at: http://www.nshd.mrc.ac.uk/data. doi: 10.5522/NSHD/Q101; doi: 10.5522/NSHD/Q102; doi: 10.5522/NSHD/Q103. Data from the 1958 birth cohort is publicly available via the UK Data Service via https://beta.ukdataservice.ac.uk/datacatalogue/series/series?id=2000032#!/abstract. Data from the 1970 birth cohort is publicly available from the UK Data Service via https://beta.ukdataservice.ac.uk/datacatalogue/series/series?id=200001. Data from the calibration sample will be made publicly available via the UK Data Service as soon as possible.

https://beta.ukdataservice.ac.uk/datacatalogue/series/series?id=2000032#!/abstract

https://beta.ukdataservice.ac.uk/datacatalogue/series/series?id=200001

## Author contributions

PP, GP and VGM formulated the research question, all authors were involved in design of the study. EG, PP and HEJ carried out the data collection. HEJ analysed the data and HEJ and PP prepared the manuscript. MR, EG, VGM and GP provided important critical revisions to the manuscript.

## Bibliography

Angold, A., Costello, E., Messer, S., Pickles, A., Winder, F., & Silver, D. (1995). The development of a short questionnaire for use in epidemiological studies of depression in children and adolescents. International Journal of Methods in Psychiatric Research, 5, 237–249.

Bell, A. (2014). Life-course and cohort trajectories of mental health in the UK, 1991-2008: a multilevel age-period-cohort analysis. Social Science & Medicine, 120, 21–30.

Blanchflower, D., & Oswald, A. (2000). Well-Being Over Time in Britain and the USA. National Bureau of Economic Research. https://doi.org/10.3386/w7487

Brazier, J. E., Harper, R., B Jones, N. M., Thomas, K. J., Usherwood, T., & Westlake, L. (1992). Validating the SF-36 health survey questionnaire: new outcome measure for primary care. British Medial Journal, 305, 160–164. https://doi.org/10.1136/bmj.305.6846.160

Chen, F., Huang, X., & MacGregor, D. (2009). Equating or linking: basic concepts and a case study. https://fliphtml5.com/xrgx/bfuj

Choi, S. W., Schalet, B., Cook, K. F., & Cella, D. (2014). Establishing a common metric for depressive symptoms: Linking the BDI-II, CES-D, and PHQ-9 to PROMIS Depression. Psychological Assessment, 26(2), 513–527. https://doi.org/10.1037/a0035768

Elliott, J., & Shepherd, P. (2006). Cohort profile: 1970 British Birth Cohort (BCS70). International Journal of Epidemiology, 35(4), 836–843. https://doi.org/10.1093/ije/dyl174

Epstein, J. F., Barker, P. R., & Kroutil, L. A. (2001). Mode Effects in Self-Reported Mental Health Data. Public Opinion Quarterly, 65(4), 529–549. https://doi.org/10.1086/323577

Farber, G., Wolpert, M., & Kemmer, D. (2020). Common Measures for Mental Health Science: Laying the Foundations. https://wellcome.org/sites/default/files/CMB-and-CMA-July-2020-pdf.pdf

Fischer, H. F., Tritt, K., Klapp, B. F., & Fliege, H. (2011). How to compare scores from different depression scales: Equating the Patient Health Questionnaire (PHQ) and the ICD-10-Symptom Rating (ISR) using Item Response Theory. International Journal of Methods in Psychiatric Research, 20(4), 203–214. https://doi.org/10.1002/mpr.350

Fischer, H. F., Wahl, I., Fliege, H., Klapp, B. F., & Rose, M. (2012). Impact of cross-calibration methods on the interpretation of a treatment comparison study using 2 depression scales. Medical Care, 50(4), 320–326. https://doi.org/10.1097/MLR.0b013e31822945b4

Furukawa, T. A., Reijnders, M., Kishimoto, S., Sakata, M., DeRubeis, R. J., Dimidjian, S., Dozois, D. A., Hegerl, U., Hollon, S. D., Jarret, R. B., Lesperance, F., Segal, Z. V, Mohr, D. C., Simons, A. D., Quilty, L. C., Reynolds III, C. F., Gentili, C., Leucht, S., Engel, R. R., & Cuijpers, P. (2019). Translating the BDI and BDI-II into the HAMD and vice versa with equipercentile linking. Epidemiology and Psychiatric Sciences, 1–13.

Goldberg, D., & Williams, P. (1988). A user’s guide to the general health questionnaire. NFER-Nelson.

Gondek, D., Bann, D., Patalay, P., Goodman, A., Richards, M., McElroy, E., & Ploubidis, G. B. (2020). Psychological distress from adolescence to early old age: Evidence from the 1946, 1958 and 1970 British birth cohorts. Psychological Medicine.

Gondek, D., Lacey, R. E., Blanchflower, D. G., & Patalay, P. (2021). How is the distribution of psychological distress changing over time? Who is driving these changes? Analysis of the 1958 and 1970 British birth cohorts. Social Psychiatry and Psychiatric Epidemiology. https://doi.org/10.1007/S00127-021-02206-6

Hambleton, R., Swaminathan, H., & Rogers, H. (1991). Fundamentals of Item Response Theory. Sage Publications ltd.

Jokela, M., Batty, G., & Kivimäki, M. (2013). Ageing and the prevalence and treatement of mental health problems. Psychological Medicine, 43, 2037–2045.

Kessler, R. C., Barker, P. R., Colpe, L. J., Epstein, J. F., Gfroerer, J. C., Hiripi, E., Howes, M. J., Normand, S. L. T., Manderscheid, R. W., Walters, E. E., & Zaslavsky, A. M. (2003). Screening for serious mental illness in the general population. Archives of General Psychiatry, 60(2), 184–189. https://doi.org/10.1001/ARCHPSYC.60.2.184

Keyes, K. M., Nicholson, R., Kinley, J., Raposo, S., Stein, M. B., Goldner, E. M., & Sareen, J. (2014). Age, Period, and Cohort Effects in Psychological Distress in the United States and Canada. Am J Epidemiol, 179(10), 1216–1227.

Kolen, M. J., & Brennan, R. L. (2014). Test Equating, Scaling and Linking (Third Edit). Springer.

Lamprianou, I. (2007). An investigation into the test equating methods used during 2006, and the potential for strengthening their validity and reliability.

Lindelow, M., Hardy, R., & Rodgers, B. (1997). Development of a scale to measure symptoms of anxiety and depression in the general UK population: the psychiatric symptom frequency scale. J Epidemiol Community Health, 51(5), 549–557.

Little, R., & Rubin, D. (2002). Statistical analyses with Missing Data (2nd Editio). Wiley.

McElroy, E., Villadsen, A., Patalay, P., Goodman, A., Richards, M., Northstone, K., Fearon, P., Tibber, M., Gondek, D., & Ploubidis, G. (2020). Harmonisation and measurement properties of mental health measures in six British cohorts.

McManus, S., Bebbington, P., Jenkins, R., & Brugha, T. (Eds.). (2016). Mental health and wellbeing in England: Adult Psychiatric Morbidity Survey 2014. NHS Digital.

MRC LHA, & NSHD. (2022). The National Survey of Health and Development. https://Www.Nshd.Mrc.Ac.Uk/.

Office for National Statistics. (2018, September 26). Personal well-being user guidance. https://www.Ons.Gov.Uk/Peoplepopulationandcommunity/Wellbeing/Methodologies/Personalwellbeingsurveyuserguide.

Piantadosi, S., Byar, D. P., & Green, S. B. (1988). The Ecological Fallacy. AMERICAN JOURNAL OF EPIDEMIOLOGY, 77(893–904).

Ploubidis, G. B., McElroy, E., & Moreira, H. C. (2019). A longitudinal examination of the measurement equivalence of mental health assessments in two british birth cohorts. Longitudinal and Life Course Studies, 10(4), 471–489. https://doi.org/10.1332/175795919X15683588979486

Ploubidis, G. B., Sullivan, A., Brown, M., & Goodman, A. (2017). Psychological distress in mid-life: evidence from the 1958 and 1970 British birth cohorts. Psychological Medicine, 47(2), 291–303. https://doi.org/10.1017/S0033291716002464

Power, C., & Elliott, J. (2006). Cohort profile: 1958 British birth cohort (National Child Development Study). International Journal of Epidemiology, 35(1), 34–41. https://doi.org/10.1093/ije/dyi183

Radloff, L. S. (2016). The CES-D Scale: A Self-Report Depression Scale for Research in the General Population. http://Dx.Doi.Org/10.1177/014662167700100306, 1(3), 385–401. https://doi.org/10.1177/014662167700100306

Rutter, M., Tizard, J., & Whitmore, K. (1970). Education, health and behaviour. Longmans.

Santor, D. A., Gregus, M., & Welch, A. (2006). Eight Decades of Measurement in Depression. Measurement: Interdisciplinary Research & Perspective, 4(3), 135–155. https://doi.org/10.1207/s15366359mea0403_1

Sellers, R., Warne, N., Pickles, A., Maughan, B., Thapar, A., & Collishaw, S. (2019). Cross-cohort change in adolescent outcomes for children with mental health problems. Journal of Child Psychology and Psychiatry and Allied Disciplines, 60(7), 813–821. https://doi.org/10.1111/jcpp.13029

Spiers, N., Brugha, T., Bebbington, P., McManus, S., Jenkins, R., & Meltzer, H. (2012). Age and birth cohort differences in depression in repeated cross-sectional surveys in England: The National Psychiatry Morbidity Surveys, 1993 to 2007. Psychological Medicine, 42(10), 2047–2055.

StataCorp. (2019). Stata Statistical Software: Release 16. StataCorp LLC.

Tennant, R., Hiller, L., Fishwick, R., Platt, S., Joseph, S., Weich, S., Parkinson, J., Secker, J., & Stewart-Brown, S. (2007). The Warwick-Edinburgh Mental Well-being Scale (WEMWBS): development and UK validation. Health and Quality of Life Outcomes, 5(1), 63. https://doi.org/10.1186/1477-7525-5-63

The Council of Chief State School Officers. (2018). A Practitioner’s Introduction to Equating.

Thomson, R. M., & Katikireddi, S. V. (2018). Mental health and the jilted generation: Using age-period-cohort analysis to assess differential trends in young people’s mental health following the Great Recession and austerity in England. Social Science and Medicine, 214, 133–143. https://doi.org/10.1016/j.socscimed.2018.08.034

UCL Centre for Longitudinal Studies. (2022a). 1958 National Child Development Study. https://Cls.Ucl.Ac.Uk/Cls-Studies/1958-National-Child-Development-Study/.

UCL Centre for Longitudinal Studies. (2022b). 1970 British Cohort Study. https://Cls.Ucl.Ac.Uk/Cls-Studies/1970-British-Cohort-Study/.

Wadsworth, M., Kuh, D., Richards, M., & Hardy, R. (2006). Cohort profile: The 1946 National Birth Cohort (MRC National Survey of Health and Development). International Journal of Epidemiology, 35(1), 49–54. https://doi.org/10.1093/ije/dyi201

White, I. R., Royston, P., & Wood, A. M. (2011). Multiple imputation using chained equations: Issues and guidance for practice. Statistics in Medicine, 30(4), 377–399. https://doi.org/10.1002/sim.4067

Whiteford, H. A., Degenhardt, L., Rehm, J., Baxter, A. J., Ferrari, A. J., Erskine, H. E., Charlson, F. J., Norman, R. E., Flaxman, A. D., Johns, N., Burstein, R., Murray, C. J. L., & Vos, T. (2013). Global burden of disease attributable to mental and substance use disorders: Findings from the Global Burden of Disease Study 2010. The Lancet, 382(9904), 1575–1586. https://doi.org/10.1016/S0140-6736(13)61611-6

Willis, S. L., Martin, M., & Rocke, C. (2010). Longitudinal perspectives on midlife development: stability and change. Eur J Ageing, 7, 131–134. https://doi.org/10.1007/s10433-010-0162-4

Wing, J., Birley, J., Cooper, J., Graham, P., & Ad, I. (1967). Reliability of a Procedure for Measuring and Classifying “Present Psychiatric State.” The British Journal of Psychiatry, 113(498), 499–515.

