## Supplemental Material for "Psychological distress across adulthood: scale-equating in three British birth cohorts"

#### Supplemental Tables

#### Supplemental Figures

### Supplemental Methods

#### *Main outcomes and measures*

Supplemental Table 1 details all psychological distress measures included in this calibration. Each measure was used at least once in at least one of the birth cohorts.

Supplemental Table 1: Measures of psychological distress used in the 1946, 1958 and 1970 British birth cohorts.

| Measure | Description | Cohorts (ages) used |
|---|---|---|
| Malaise-9 questionnaire | Nine items covering emotional disturbance and associated somatic symptoms. No timescale indicated, binary (yes/no) response options.<br>Main symptoms: low mood, fatigue, stress, worry, general anxiety, irritability, panic<br>Total possible score range: 0-9. Cut-off ≥ 4. | 1958 (age 23,33,42,50)<br>1970 (age 26,30,34,42,46) |
| General Health Questionnaire (GHQ-12) | 12 items from the original General Health Questionnaire aimed at detecting psychiatric disorders in the community. Responses recorded on a 4-point Likert scale. Timeline: the past few weeks.<br>Main symptoms: low mood, stress, sleep problems, loss of interest, concentration problems, indecision, worthlessness<br>Total possible score range: 0-36. Cut-off ≥ 12. | 1958 (age 42)<br>1970 (age 30) |
| General Health Questionnaire (GHQ-28) | 28 items from the original General Health Questionnaire aimed at detecting psychiatric disorders in the community. Responses recorded on a binary scale. Timeline: the past few weeks.<br>Main symptoms: fatigue, stress, sleep problems, irritability, panic, hopelessness, loss of interest, health anxiety, indecision, somatic complaints, worthlessness, loss of motivation, suicidal ideation.<br>Total possible score range: 0-28. Cut-off ≥ 9. | 1946 (age 53, 63, 69) |
| Present State Examination (PSE) | Nine items designed to record psychiatric symptoms. Responses recorded on a 3-point Likert scale.<br>Main symptoms: low mood, fatigue, sleep problems, worry, general anxiety, irritability, panic, hopelessness, loss of interest, concentration problems, appetite disturbance, health anxiety, restlessness, social phobia, indecision, somatic complaints, worthlessness, suicidal ideation.<br>Total possible score range: 0-18. | 1946 (age 36) |
| Psychiatric Symptom Frequency Scale (PSF) | 18 items designed to assess symptoms of anxiety and depression experienced over the past year in the general population. Responses recorded on a 6-point Likert scale.<br>**Main symptoms:** low mood, fatigue, stress, sleep problems, general anxiety, panic, hopelessness, loss of interest, concentration problems, loss of functionality, appetite disturbance, health anxiety, restlessness, loss of motivation, suicidal ideation.<br>Total possible score range: 0-90. Cut-off ≥ 23. | 1946 (age 43) |

#### *Assumptions underlying equipercentile linking*

##### 1. Symmetry

The function used to transform scores from all measures into the GHQ-12 or Malaise-9 is the inverse of the function to transform GHQ-12/Malaise-9 into other measures, as these are based on percentile scorings. See Table 1 and Supplemental Table 5.

2. Content equivalence

Based on a previous harmonisation project, we are confident that all measures included here capture the phenomenon of psychological distress to a sufficiently similar extent.

3. Similar and acceptable reliability of scores

Using our calibration sample, we computed Cronbach's alpha for all measures included in this calibration project (Supplemental Table 2). These varied between 0.86 for the Malaise-9 to 0.97 for the PSF, and we consider these to be both acceptable and sufficiently similar.

Supplemental Table 2:Cronbach's alpha for each measure

| Measure | Cronbach's alpha |
|---|---|
| Malaise-9 | 0.86 |
| GHQ-12 | 0.91 |
| GHQ-28 | 0.94 |
| PSE | 0.88 |
| PSF | 0.97 |

4. Equity

Whilst we didn't formally test for equity as part of this study, we did randomise the order of administration of our mental health measures to prevent order effects.

5. Observed score equating

There is no gold standard measure of psychological distress nor any 'true' score, so this assumption has been met.

6. Group invariance

Measurement invariance between younger (up to 40) and older (41 or over) participants was tested for all measures that were applied to both these age groups (Supplemental Table 3 below). For the GHQ-12, there were mixed findings both the Standardised Root Mean Square Residual (SRMR) and the Comparative Fit Index (CFI) indicated invariance at the configural, the metric and scalar level.. For the Root Mean Square Error of Approximation (RMSEA) and SRMR, a score closest to zero indicates good model fit, whereas for CFI it is closest to one. Rules of thumb for model comparison are as follows: for RMSEA invariance is met if a change is smaller than 0.015, for SRMR if it is smaller than 0.03 and for CFI if it is smaller than 0.01 (Chen, 2007). Across both the GHQ-28 and the Malaise-9 all three indicators suggested scalar invariance. We are therefore satisfied that the measurement invariance assumption is met and the tests can be considered equivalent across the two age groups.

**Supplemental Table 3: Group invariance for GHQ-12, GHQ-28 and Malaise-9**

| | Configural | Metric | Scalar |
|---|---|---|---|
| GHQ-12 | | | |
| RMSEA[1] | 0.132 | 0.125 | 0.107 |
| SRMR[2] | 0.047 | 0.050 | 0.050 |
| CFI[3] | 0.953 | 0.953 | 0.959 |
| GHQ-28 | | | |
| RMSEA[1] | 0.069 | Not applicable | 0.069 |
| SRMR[2] | 0.069 | | 0.070 |
| CFI[3] | 0.950 | | 0.950 |
| Malaise-9 | | | |
| RMSEA[1] | 0.058 | Not applicable | 0.057 |
| SRMR[2] | 0.048 | | 0.049 |
| CFI[3] | 0.989 | | 0.988 |

[1] Root Mean Square Error of Approximation
[2] Standardised Root Mean Square Residual
[3] Comparative Fit Index

#### *Multiple imputation*

The multiple imputation framework is summarised in Supplemental Table 1 below.

Supplemental Table 4: Imputation framework

| | NSHD 1946 | | NCDS 1958 | | BCS 1970 | |
|---|---|---|---|---|---|---|
| **Age-group** | Available[1] | Imputed[2] | Available[1] | Imputed[2] | Available[1] | Imputed[2] |
| 23-30 | -- | -- | Malaise-9 | GHQ-12 | Malaise-9<br>GHQ-12 | |
| 31-40 | PSE | Malaise-9<br>GHQ-12 | Malaise-9 | GHQ-12 | Malaise-9 | GHQ-12 |
| 41-50 | PSF | Malaise-9<br>GHQ-12 | Malaise-9<br>GHQ-12 | | Malaise-9 | GHQ-12 |
| 51-60 | GHQ-28 | Malaise-9<br>GHQ-12 | -- | -- | -- | -- |
| 61-70 | GHQ-28 | Malaise-9<br>GHQ-12 | -- | -- | -- | -- |

[1] Data available in both the calibration sample and the birth cohort
[2] Data to be imputed in the cohort sample, available from the calibration sample

#### Supplemental Results

The distribution of sex and level of education for each of our four samples, as well as the questionnaires administered at each age can be found in Supplemental Table 5 below.

Supplemental Table 5: Description of analysis samples

| Sample | n | Men (%) | Level of education | | Measures (age administered) |
|---|---|---|---|---|---|
| Calibration[1] | 5,800 | 3,000 (51.7%) | No qualifications | 311 (5.4%) | GHQ-28 (all |
| | | | GSCE | 2,000 (34.5%) | GHQ-12 (all) |
| | | | A-levels | 1,483 (25.6%) | Malaise-9 (all) |
| | | | Higher qualifications | 2,006 (34.6%) | PSE (31-40) |
| | | | Missing | 0 (0.0%) | PSF (41-50) |
| 1946 cohort | 5,362 | 2,815 (52.5%) | No qualifications | 1,579 (29.5%) | GHQ-28 (53, 63, 69) |
| | | | GSCE | 1,393 (26.0%) | PSE (36) |
| | | | A-levels | 1,089 (20.3%) | PSF (43) |
| | | | Higher qualifications | 558 (10.4%) | |
| | | | Missing | 743 (13.9%) | |
| 1958 cohort | 14,814 | 7,509 (50.7%) | No qualifications | 1,910 (12.9%) | Malaise-9 (23, 33, 42, 50) |
| | | | GSCE | 4,739 (32.0%) | GHQ-12 (42) |
| | | | A-levels | 1,226 (8.3%) | |
| | | | Higher qualifications | 1,908 (12.9) | |
| | | | Missing | 5,031 (34.0%) | |
| 1970 cohort | 13,739 | 6,828 (49.7%) | No qualifications | 3,792 (27.6%) | Malaise-9 (26, 30, 34, 42, 46) |
| | | | GSCE | 4,782 (34.8%) | GHQ-12 (30) |
| | | | A-levels | 1,008 (7.3%) | |
| | | | Higher qualifications | 2,369 (17.2%) | |
| | | | Missing | 1,788 (13.0%) | |

Distribution across age-groups was as follows:
23-30: 1,167
31-40: 1,170
41-50: 1,163
51-60: 1,163
61-70: 1,135

The number of respondents, means and standard deviations for each measure in the calibration sample can be found in Supplemental Table 6 below.

Supplemental Table 6: Number of respondents, means and standard deviations for each measure in the calibration sample

| Measure | Number of respondents (%) | Mean (SD) |
|---|---|---|
| Malaise-9 | 5,665 (97.7%) | 3.16 (2.80) |
| General Health Questionnaire-12 | 5,641 (97.3%) | 14.29 (6.36) |
| General Health Questionnaire-28 | 5,505 (94.9%) | 28.71 (16.51) |
| Present State Examination[1] | 1,142 (97.6%) | 7.22 (5.43) |
| Psychiatric Symptom Frequency Scale[2] | 1,116 (96.0%) | 31.77 (21.82) |

[1] PSE is only administered in those aged 31-40 (n=1,170)
[2] PSF is only administered in those aged 41-50 (n=1.163)

Correlations between all measures administered in the calibration sample are detailed in Supplemental Table 7 below.

**Supplemental Table 7: Correlations between all measures administered in the calibration sample**

| | Malaise-9 | GHQ-12 | GHQ-28 | PSE | PSF |
|---|---|---|---|---|---|
| Malaise-9 | -- | -- | -- | -- | -- |
| GHQ-12 | 0.68 | -- | -- | -- | -- |
| GHQ-28 | 0.74 | 0.91 | -- | -- | -- |
| PSE | 0.73 | 0.71 | 0.76 | -- | -- |
| PSF | 0.76 | 0.70 | 0.76 | --[1] | -- |

[1] No correlation could be computed between PSE and PSF as these were administered in different age groups.

The distribution of all total scores calibrated against the GHQ-12 using a equipercentile linking approach can be found in Supplemental Figure 1, alongside the distribution of total scores derived using a multiple imputation approach (Supplemental Figure 2) and the distribution of total scores of all original measures included in all birth cohorts (see Supplemental Table 1) in Supplemental Figure 3.

Supplemental Figure 1: Histograms of total scores calibrated against the GHQ-12 using the equipercentile linking method.

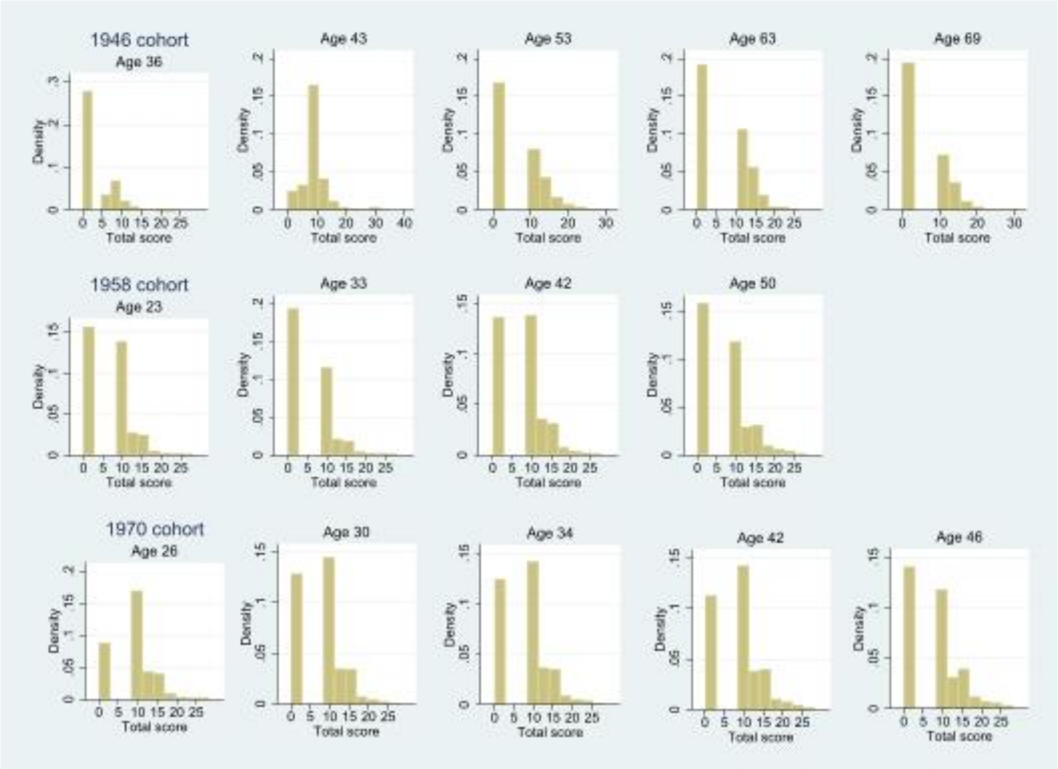

Supplemental Figure 2: Histograms of total scores calibrated against the GHQ-12 using the multiple imputation method.

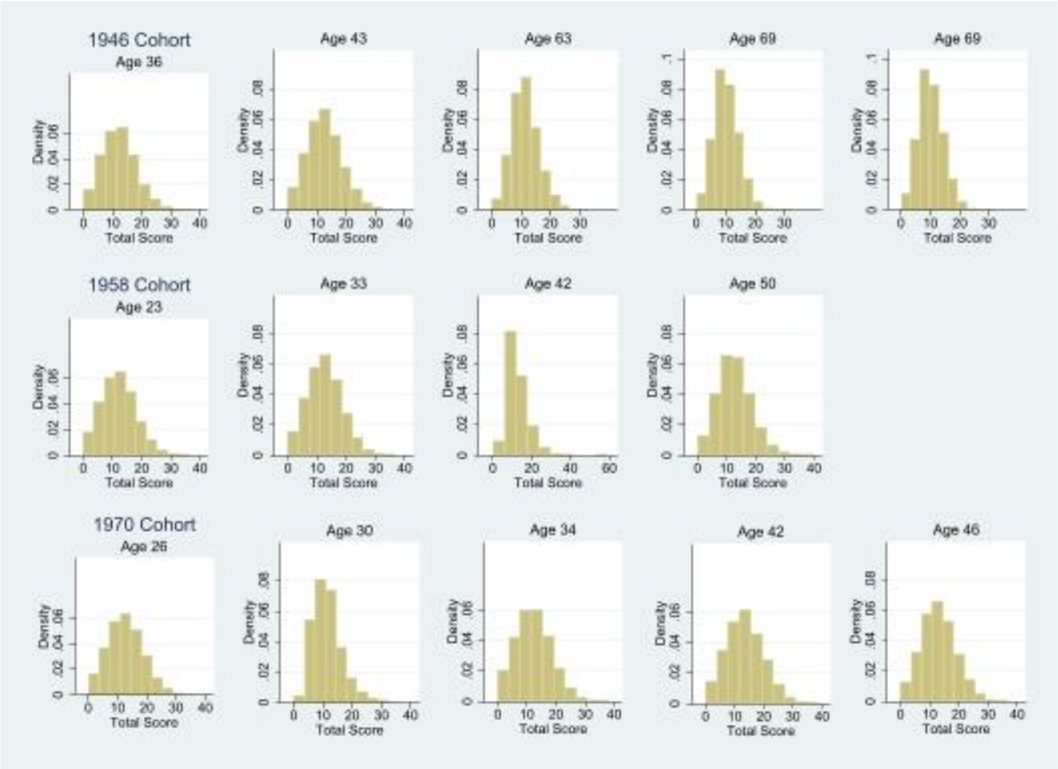

Supplemental Figure 3: Histograms of total scores of original measures

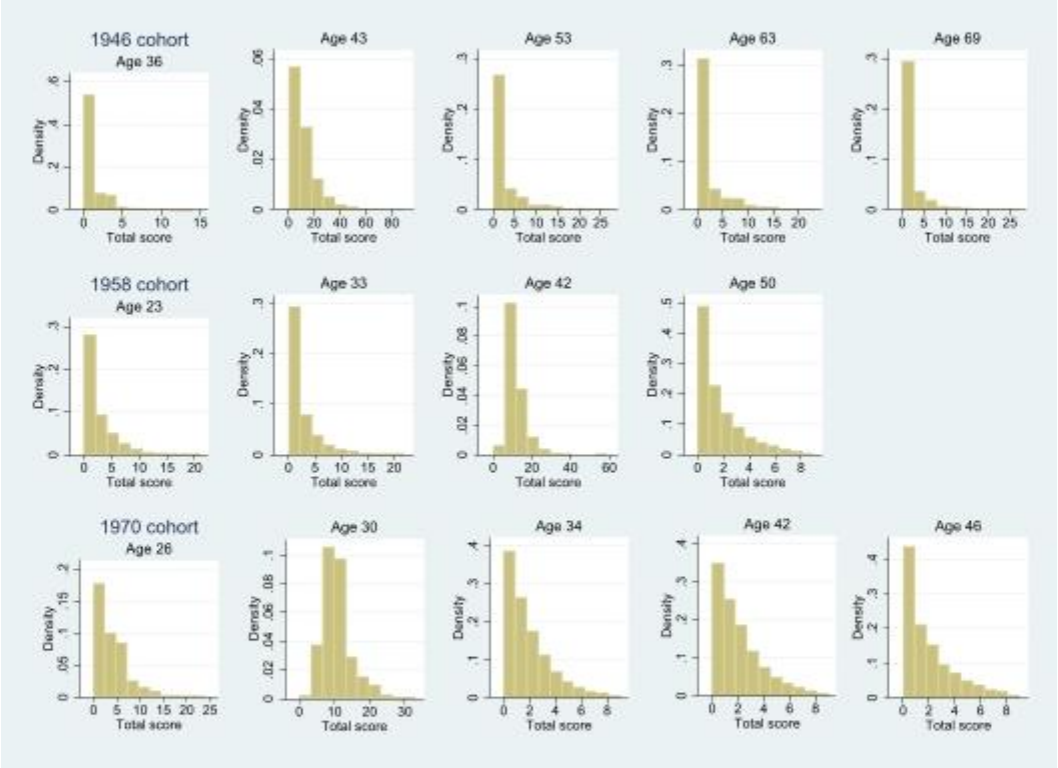

##### *Calibration against the Malaise-9*

*Equipercentile linking*

Scores calibrated to Malaise-9 are detailed in Supplemental Table 7. Means and corresponding standard deviations across the life course in each of the three cohorts are detailed in Supplemental Table 8. Corresponding calibrated cut-off scores to the Malaise-9 were 9 for Malaise-24, 14 for GHQ-12, 6 for GHQ-28, 8 for PSE and 32 for PSF (Supplemental Table 7). Psychological distress peaked in the 1946 birth cohort at age 53 (mean: 1.69, SD: 1.98) before gradually declining to 1.28 (SD:1.85) at age 69. In the 1958 birth cohort this peak was earlier and higher, at age 42 (mean: 2.17, SD: 2.36), with psychological distress being lower again at age 50 (mean: 1.49, SD: 1.94). In the 1970 cohort psychological distress peaked at age 30 (mean: 2.00, SD: 2.19) followed by a non-linear decline to age 46 (mean: 1.77, SD: 2.12). Across the life course and across cohorts, distributions tended to be positively skewed (Supplemental Figure 4).

Supplemental Table 8: Calibrated scores and cut-offs against the Malaise-9

| | Calibrated equivalent scores per questionnaire | | | | |
|---|---|---|---|---|---|
| Malaise-9 score | Malaise-24 | GHQ-12 | GHQ-28 | PSE | PSF |
| 0 | 1,2 | 0-7 | 0 | 0,1 | 1-4 |
| 1 | 2,3 | 8-10 | | 2-4 | 5-15 |
| 2 | 4-7 | 11,12 | 1,2 | 5,6 | 16-21 |
| 3 | 7 | 13 | 3,4 | 7 | 22-28 |
| 4 | 8,**9** | **14** | 5-7 (**6**) | **8** | 29-36 (**32**) |
| 5 | 10,11 | 15,16 | 8-10 | 9 | 37-42 |
| 6 | 12,13 | 17,18 | 11-13 | 10,11 | 43-48 |
| 7 | 14,15 | 19-21 | 14-17 | 12 | 49-55 |
| 8 | 16-18 | 22-25 | 18-21 | 13,14 | 56-66 |
| 9 | 19-24 | 26-35 | 22-28 | 15-18 | 67-90 |

Equivalent cut-off scores are in **bold**

Supplemental Table 9: Means and standard deviations of measures calibrated against the Malaise-9 across the life course

| | | Multiple imputation | Equipercentile linking | Original measure |
|---|---|---|---|---|
| Cohort | Age | Mean (SD) | Mean (SD) | Mean (SD) |
| **1946** | 36 | 2.11 (0.15) | 0.32 (0.71) | |
| | 43 | 2.11 (0.12) | 1.22 (1.42) | |
| | 53 | 2.29 (0.12) | 1.69 (2.17) | |
| | 64 | 2.07 (0.10) | 1.64 (1.98) | |
| | 69 | 1.28 (0.10) | 1.28 (1.85) | |
| **1958** | 23 | 1.29 (0.01) | 1.24 (1.27) | |
| | 33 | 1.07 (0.01) | 1.01 (1.55) | |
| | 42 | 1.60 (0.02) | 2.17 (2.36) | 1.52 (1.78) |
| | 50 | 1.59 (0.02) | 1.49 (1.94) | |
| **1970** | 26 | 1.87 (0.02) | 1.82 (1.75) | |
| | 30 | 1.60 (0.02) | 2.00 (2.19) | 1.56 (1.76) |
| | 34 | 1.77 (0.02) | 1.68 (1.90) | |
| | 42 | 1.95 (0.02) | 1.85 (1.99) | |
| | 46 | 1.90 (0.02) | 1.77 (2.12) | |

Distribution of above-threshold prevalence is detailed in Supplemental Figure 6. For our calibrated cut-off scores in the 1946 cohort, prevalence of psychological distress increased to a peak at age 53 (at 16.4%) before declining again. In the 1958 cohort prevalence peaked at age 42 at 22.0% and in the 1970 cohort it peaked at age 34 at 19.8%. Using the equipercentile linking method (where total scores were calibrated before a cut-off was applied), a similar pattern was observed. Prevalence of psychological distress in the 1946 cohort peaks at age 53 at 19.4%, in the 1958 birth cohort at age 42 at 22.0% and in the 1970 birth cohort at age 34 at 19.8%.

Supplemental Figure 4: Histograms of mental health measures calibrated against the Malaise-9 using the equipercentile linking method

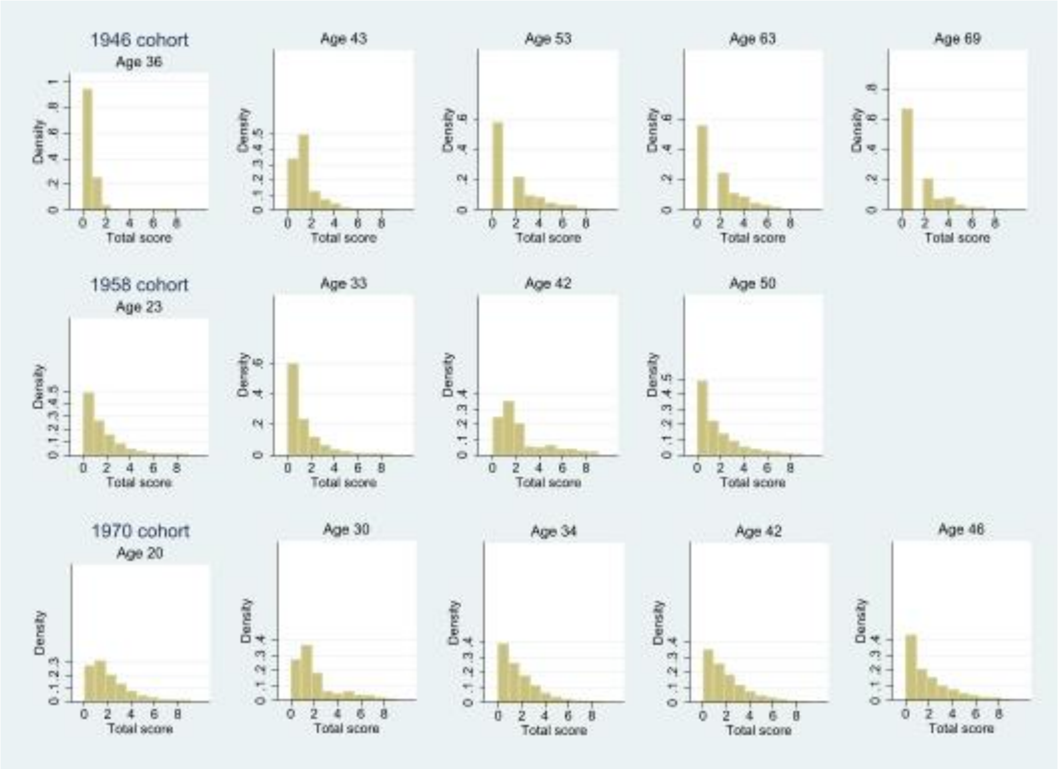

*Multiple imputation*

Means and standard deviations of psychological distress across the life course based on multiple imputation are detailed in Supplemental Table 8 above. In the 1946 birth cohort, psychological distress peaked at age 53 with a mean of 2.29 and SD of 0.12 before declining to 2.07 (SD: 0.10) at age 69. In the 1958 birth cohort, there was a plateau from age 42 onwards (age 42 mean:1.60, SD: 0.02; age 50 mean: 1.59, SD: 0.02) and the mean score remained relatively stable in the 1970 birth cohort, with a small peak at age 42 (mean:1.95, SD:0.02). The distribution of total scores based on multiple imputation is detailed in Supplemental Figure 5 below. At most sweeps, the distribution was positively skewed.

Supplemental Figure 5:Histograms of total scores of mental health measures calibrated against the Malaise-9 using the multiple imputation method.

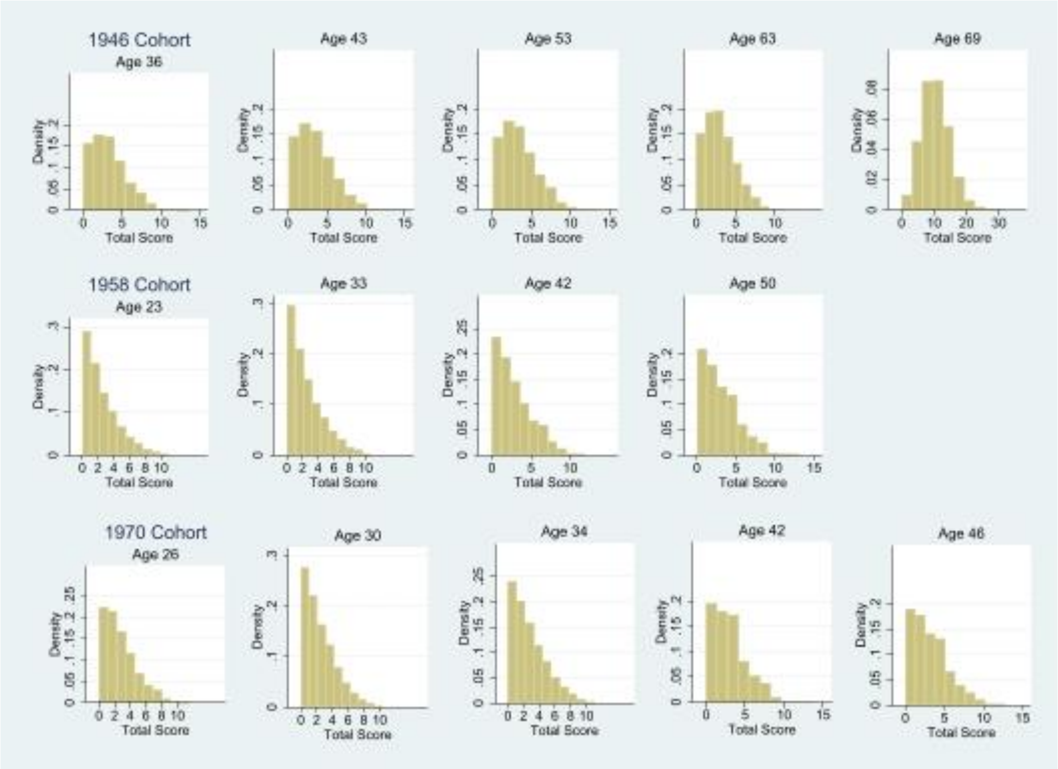

Prevalence estimates are included in Supplemental Figure 6 below. In the 1946 birth cohort, prevalence of psychological distress peaked at age 53 (20.5%). In both the 1958 and 1970 birth cohorts, we observed a gradual increase in the prevalence of psychological distress across the life course, with prevalence peaking at age 50 in the 1958 cohort (14.8%) and at age 46 in the 1970 birth cohort (19.3%).

Supplemental Figure 6: Prevalence of psychological distress across the three birth cohorts (calibrated against the Malaise-9)

Supplemental Figure 6A: The 1946 birth cohort

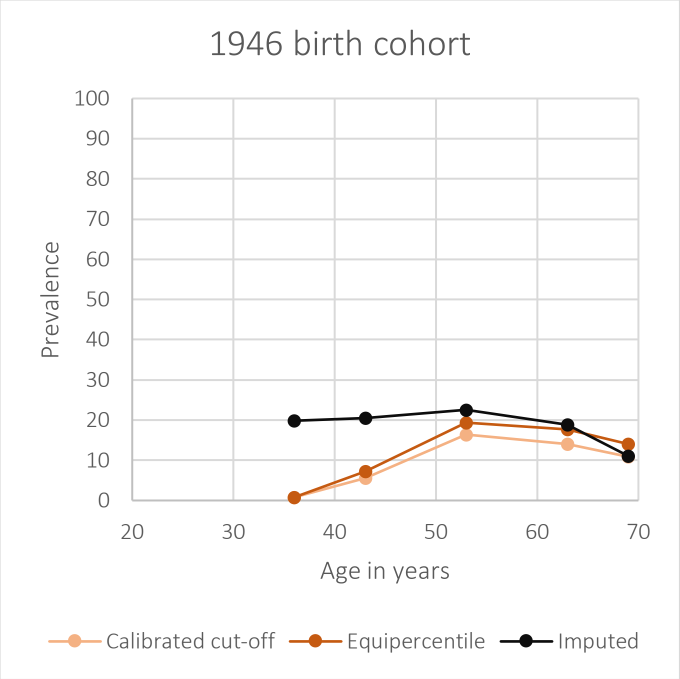

Supplemental Figure 6B: The 1958 birth cohort

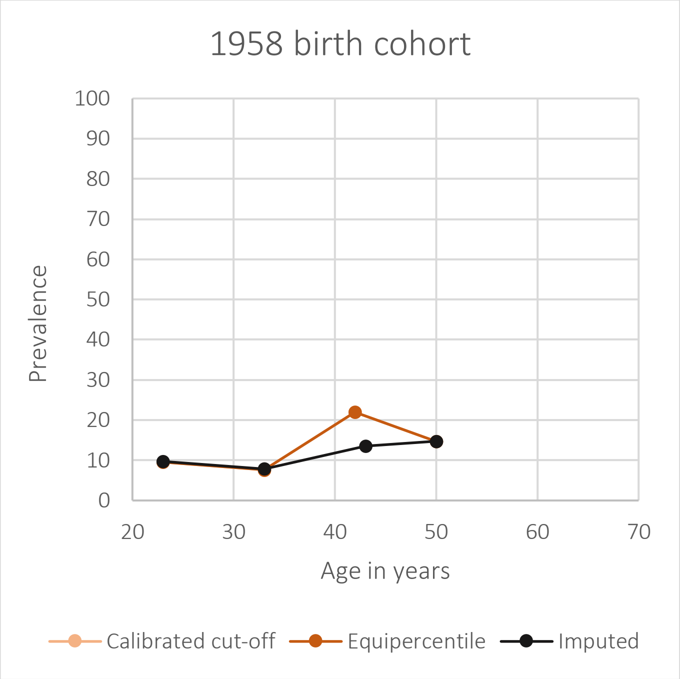

Supplemental Figure 6C: The 1970 birth cohort

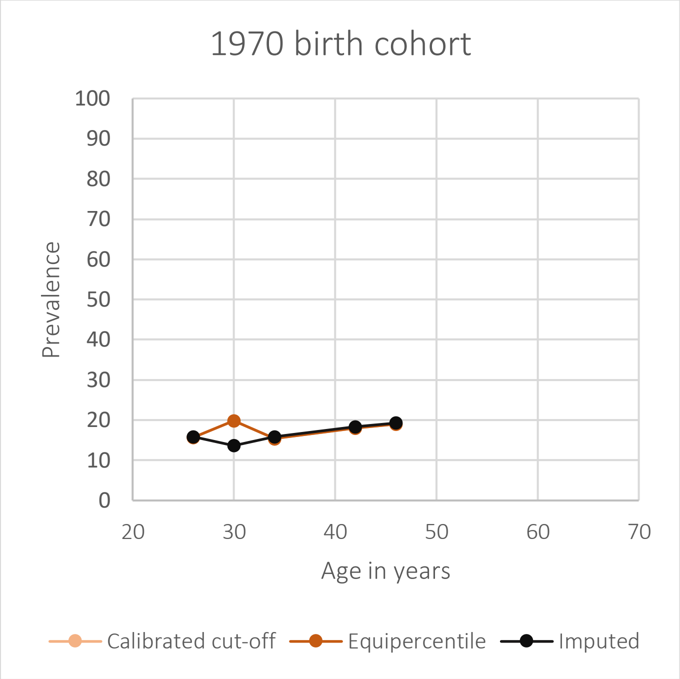

In the 1958 and 1970 birth cohort, the Malaise-9 questionnaire was available throughout the life-course, and as such there was no difference in the various calibration methods. The data points (age 42 in the 1958 birth cohorts, and age 34 in the 1970) where divergence is observed is the only sweep where the GHQ-12 was also available, and has been calibrated against the Malaise-9. As the cut-off scores for both the equipercentile linking and the calibrated cut-off scores were identical, the calibrated cut-off method is not visible on the graphs.

*Sensitivity analyses*

Supplemental Table 9 below details the mean scores (where possible) and prevalence of psychological distress using the two sweeps where both the GHQ-12 and Malaise-9 were administered, age 42 in the 1958 cohort and at age 30 in the 1970 cohort, allowing comparison of prevalence resulting from the original measure and all three calibration methods.

Across both measures and birth cohorts, the multiple imputation approach approximates original mean scores and prevalences most closely. For the Malaise-9 in the 1958 cohort both a calibrated cut-off and equipercentile linking approach overestimated prevalence. For the GHQ-12 at this sweep, the calibrated cut-off approximated the original prevalence, whereas the equipercentile linking method underestimated the mean and prevalence. For the Malaise-9 in the 1970 cohort, both the calibrated cut-off and equipercentile linking method overestimated means and prevalence. For the GHQ-12 at this sweep, the calibrated cut-off in an overestimation, whereas the equipercentile linking approach resulted in underestimating psychological distress.

**Supplemental Table 10: Mean and prevalence of psychological distress using original data and three calibration methods at two sweeps.**

| | Original | Calibrated-cut-off | Equipercentile linking | Multiple imputation |
|---|---|---|---|---|
| 1958 age 42 | | | | |
| *GHQ-12* | | | | |
| Mean (SD) | 11.06 (4.79) | n/a | 7.26 (6.23) | 11.19 (0.05) |
| Prevalence | 36.1% | 38.5% | 23.1% | 38.5% |
| *Malaise-9* | | | | |
| Mean (SD) | 1.52 (1.78) | n/a | 2.17 (2.36) | 1.60 (0.02) |
| Prevalence | 13.0% | 22.0% | 22.0% | 13.0% |
| 1970 age 30 | | | | |
| *GHQ-12* | | | | |
| Mean (SD) | 10.67 (4.52) | n/a | 7.48 (6.11) | 10.74 (0.04) |
| Prevalence | 32.9% | 39.8% | 23.5% | 34.5% |
| *Malaise-9* | | | | |
| Mean (SD) | 1.56 (1.76) | n/a | 2.00 (2.19) | 1.60 (0.02) |
| Prevalence | 13.7% | 19.8% | 19.8% | 13.7% |

#### Supplemental bibliography

Chen, F. F. (2007). Sensitivity of goodness of fit indexes to lack of measurement invariance. *Structural Equation Modeling*, *14*(3), 464–504. https://doi.org/10.1080/10705510701301834
